# Anti-cardiolipin and other anti-phospholipid antibodies in critically ill COVID-19 positive and negative patients

**DOI:** 10.1101/2021.02.19.21252113

**Authors:** Uriel Trahtemberg, Robert Rottapel, Claudia C dos Santos, Arthur S. Slutsky, Andrew J Baker, Marvin J Fritzler on behalf of the COVID19 Longitudinal Biomarkers of Lung Injury (COLOBILI) group.

## Abstract

**Background:** Reports of severe COVID-19 being associated with thrombosis, anti-phospholipid antibodies (APLA), anti-phospholipid syndrome (APS) have yielded disparate conclusions. Studies comparing COVID-19 patients with contemporaneous controls of similar severity are lacking.

**Methods:** 22 COVID^+^ and 20 COVID^−^ patients with respiratory failure admitted to intensive care were studied longitudinally. Demographic and clinical data were obtained from the day of admission. APLA testing included anti-cardiolipin (aCL), anti-β2glycoprotien 1 (β2GP1), anti-domain 1 beta2 glycoprotein 1 (β2GP1) and anti-phosphatidyl serine/prothrombin complex (PS/PT). Anti-nuclear antibodies (ANA) were detected by immunofluorescence and antibodies to cytokines by a commercially available multiplexed array. ANOVA was used for continuous variables and Fisher’s exact test was used for categorical variables with α=0.05 and the false discovery rate at q=0.05.

**Results:** APLA were predominantly IgG aCL (48%) followed by IgM (21%) in all patients, with a tendency toward higher frequency among the COVID^+^. aCL was not associated with surrogate markers of thrombosis but IgG aCL was strongly associated with worse disease severity and higher ANA titers regardless of COVID-19 status. An association between aCL and anti-cytokine autoantibodies tended to be higher among the COVID^+^.

**Conclusions:** Positive APLA serology was associated with more severe disease regardless of COVID-19 status.

**KEY MESSAGES:** *What is already known about this subject?:*

- COVID-19 is associated with coagulopathy and high morbidity and mortality.
- COVID-19 shares some of these clinical features with anti-phospholipid syndrome.
- Reports of an association of anti-phospholipid antibodies with high risk COVID-19 have yielded disparate conclusions, but they lacked longitudinal follow up and control groups of similar severity.

*What does this study add?:*

- Anti-phospholipid syndrome serology assessed longitudinally was predominantly anticardiolipin IgG autoantibodies, in 48% of patients.
- Anticardiolipin serology was associated with worse disease severity in both COVID-19 positive and negative patients.

*How might this impact on clinical practice or future developments?:*

- The use of anti-phospholipid antibodies tests in the COVID-19 clinical setting needs to be taken in context; whereas they are associated with more serve disease, they do not discriminate between COVID-19 positive and negative patients.

## INTRODUCTION

Anti-phospholipid antibodies (APLA) are biomarkers of a spectrum of clinical features observed in anti-phospholipid syndrome (APS) ^1^. Features of APS include venous and arterial thrombosis involving multiple organs and having various presentations ^1^. APLA that are components of APS criteria include IgG and/or IgM anti-cardiolipin (aCL), -β2-glycoprotein1 (GP1), and the “lupus anticoagulant” (LAC)^2^. Other non-criteria APLA such as anti-phosphatidylserine/prothrombin (PS/PT) complex, -PT, and -domain 1 of β2-GP1 have also found a diagnostic niche in APS^3 4^.

One of the salient features of COVID-19 is the development of thrombotic events associated with severe morbidity and mortality ^5-8^. In the context of systemic inflammation and dysregulated immunity^9^, some reports have linked APLA to these thromboses ^10 11^, severe COVID-19^5 12^ and release of neutrophil extracellular traps^5^. However, APLA are also described in a variety of other infectious diseases^13^ and critically ill patients have high rates of thromboembolism that were not linked to APS or APLA^14^. Therefore, the association of COVID-19 with APLA and their potential pathogenic role ^15^ has not been clearly demonstrated due to the lack of contemporaneous, COVID-19 negative controls. Here we compare the prevalence and clinical correlations of APLA in patients with severe COVID-19 as compared to contemporaneous non-COVID19 patients with similar clinical characteristics.

## METHODS

This is an observational cohort study of patients admitted to St. Michael’s Hospital (Toronto, ON, Canada), as approved by the Research Ethics Board (REB# 20-078). Informed consent was obtained from all patients or their legal surrogates. Inclusion criteria were age ≥18 years, admission to intensive care unit (ICU) with acute respiratory failure. Exclusion criteria were inability to ascertain the primary outcome or obtain a baseline blood sample, and SARS-CoV2 infection in the 4 weeks prior to admission. COVID-19 status was determined with PCR of nasopharyngeal swabs and/or endotracheal aspirates. Follow-up was 3 months post-ICU admission or hospital discharge. Primary outcome was death in the ICU. Secondary outcomes were in hospital-death, ICU utilization metrics, organ dysfunction measures and severity scores. Clinical data and serum samples were collected longitudinally at days 0, 1, 3, 5, 7 and 10; after day 10 or ICU discharge. Anti-CL, anti-β2-GP1 and anti-PS/PT were tested for IgG and IgM, as well as IgG anti-domain 1 β2-GP1; all by ELISA or chemiluminescence (Inova Diagnostics, San Diego, CA USA). ANOVA was used for continuous variables and Fisher’s exact test was used for categorical variables at α=0.05, followed by a false discovery rate adjustment at q=0.05. Detailed methods are available (online supplement), including methods for detection of anti-nuclear autoantibodies (ANA) by HEp-2 immunofluorescence assay (IFA) (Inova Diagnostics, San Diego, CA USA), and antigen-specific autoantibodies (TheraDiag, Paris, France) and anti-cytokine autoantibodies (Millipore, Oakville, ON, Canada) using addressable laser bead immunoassays.

## RESULTS

The demographic and clinical parameters of 22 COVID-19 positive (COVID^+^) and 20 COVID-19 negative (COVID^-^) patients (Table 1) included an average of 14.1 day stays in ICU and 31% mortality, but no statistically significant differences between the two cohorts, including the lack of significant differences in the number of thrombotic events requiring therapeutic anticoagulation, platelet counts, or platelet counts normalized to the neutrophil counts (to index for severity) (Table 1). None of the patients had a history of antecedent APS, systemic lupus erythematosus (SLE) or other conditions associated with APS, nor were there significant differences in other past medical history between COVID^+^ and COVID^-^ patients (supplemental table 1).

**Table 1:**
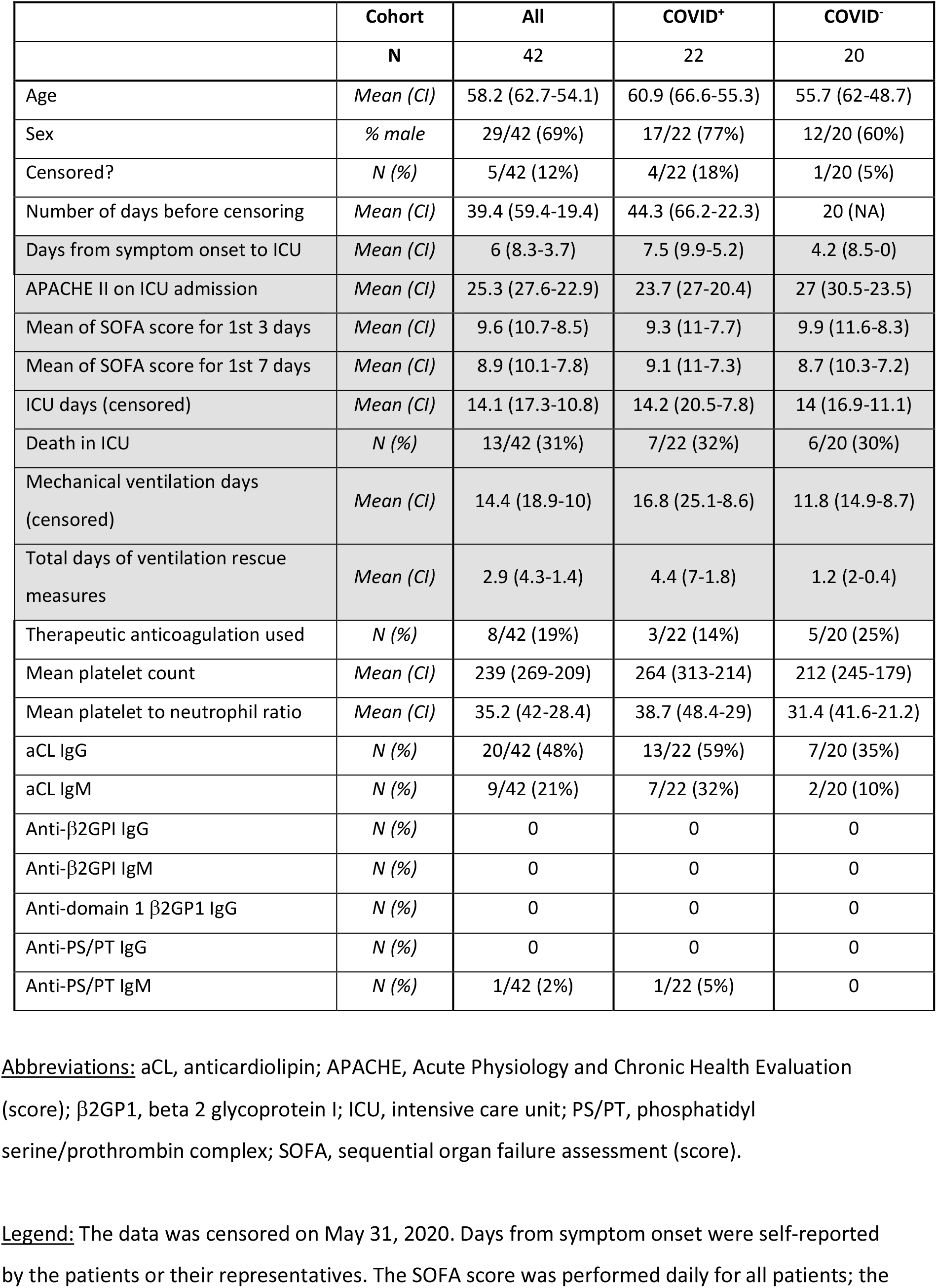
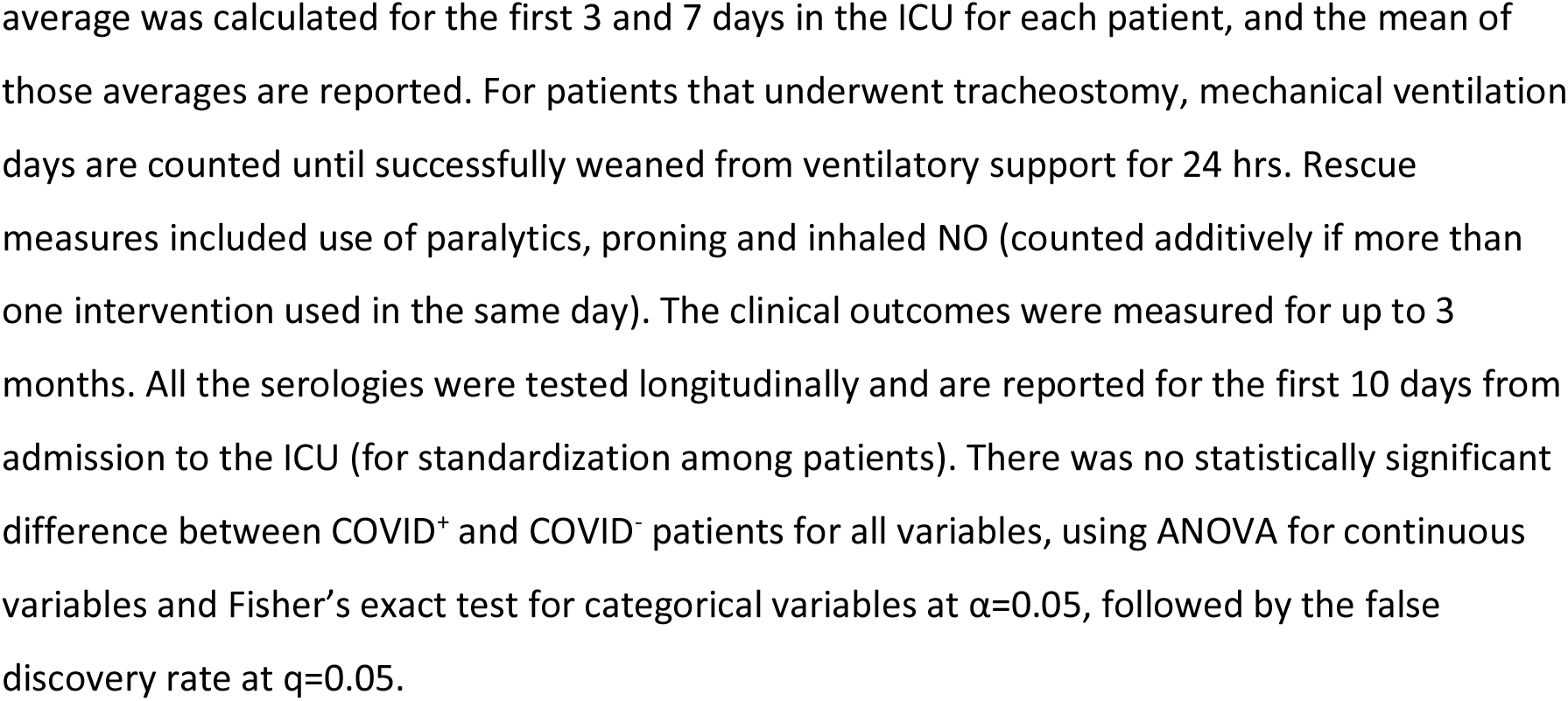
Patient Demographic, Clinical and Autoantibody Status

|  | Cohort | All | COVID<sup>+</sup> | COVID<sup>-</sup> |
| --- | --- | --- | --- | --- |
|  | N | 42 | 22 | 20 |
| Age | Mean (CI) | 58.2 (62.7-54.1) | 60.9 (66.6-55.3) | 55.7 (62-48.7) |
| Sex | % male | 29/42 (69%) | 17/22 (77%) | 12/20 (60%) |
| Censored? | N (%) | 5/42 (12%) | 4/22 (18%) | 1/20 (5%) |
| Number of days before censoring | Mean (CI) | 39.4 (59.4-19.4) | 44.3 (66.2-22.3) | 20 (NA) |
| Days from symptom onset to ICU | Mean (CI) | 6 (8.3-3.7) | 7.5 (9.9-5.2) | 4.2 (8.5-0) |
| APACHE II on ICU admission | Mean (CI) | 25.3 (27.6-22.9) | 23.7 (27-20.4) | 27 (30.5-23.5) |
| Mean of SOFA score for 1st 3 days | Mean (CI) | 9.6 (10.7-8.5) | 9.3 (11-7.7) | 9.9 (11.6-8.3) |
| Mean of SOFA score for 1st 7 days | Mean (CI) | 8.9 (10.1-7.8) | 9.1 (11-7.3) | 8.7 (10.3-7.2) |
| ICU days (censored) | Mean (CI) | 14.1 (17.3-10.8) | 14.2 (20.5-7.8) | 14 (16.9-11.1) |
| Death in ICU | N (%) | 13/42 (31%) | 7/22 (32%) | 6/20 (30%) |
| Mechanical ventilation days (censored) | Mean (CI) | 14.4 (18.9-10) | 16.8 (25.1-8.6) | 11.8 (14.9-8.7) |
| Total days of ventilation rescue measures | Mean (CI) | 2.9 (4.3-1.4) | 4.4 (7-1.8) | 1.2 (2-0.4) |
| Therapeutic anticoagulation used | N (%) | 8/42 (19%) | 3/22 (14%) | 5/20 (25%) |
| Mean platelet count | Mean (CI) | 239 (269-209) | 264 (313-214) | 212 (245-179) |
| Mean platelet to neutrophil ratio | Mean (CI) | 35.2 (42-28.4) | 38.7 (48.4-29) | 31.4 (41.6-21.2) |
| aCL IgG | N (%) | 20/42 (48%) | 13/22 (59%) | 7/20 (35%) |
| aCL IgM | N (%) | 9/42 (21%) | 7/22 (32%) | 2/20 (10%) |
| Anti-β2GPI IgG | N (%) | 0 | 0 | 0 |
| Anti-β2GPI IgM | N (%) | 0 | 0 | 0 |
| Anti-domain 1 β2GP1 IgG | N (%) | 0 | 0 | 0 |
| Anti-PS/PT IgG | N (%) | 0 | 0 | 0 |
| Anti-PS/PT IgM | N (%) | 1/42 (2%) | 1/22 (5%) | 0 |

Abbreviations: aCL, anticardiolipin; APACHE, Acute Physiology and Chronic Health Evaluation
(score); β2GP1, beta 2 glycoprotein I; ICU, intensive care unit; PS/PT, phosphatidyl
serine/prothrombin complex; SOFA, sequential organ failure assessment (score).
Legend: The data was censored on May 31, 2020. Days from symptom onset were self-reported
by the patients or their representatives. The SOFA score was performed daily for all patients; the
average was calculated for the first 3 and 7 days in the ICU for each patient, and the mean of those averages are reported. For patients that underwent tracheostomy, mechanical ventilation days are counted until successfully weaned from ventilatory support for 24 hrs. Rescue measures included use of paralytics, proning and inhaled NO (counted additively if more than one intervention used in the same day). The clinical outcomes were measured for up to 3 months. All the serologies were tested longitudinally and are reported for the first 10 days from admission to the ICU (for standardization among patients). There was no statistically significant difference between COVID<sup>+</sup> and COVID<sup>-</sup> patients for all variables, using ANOVA for continuous variables and Fisher's exact test for categorical variables at $\alpha=0.05$ , followed by the false discovery rate at $q=0.05$ .

### Frequency, development and distribution of aCL

Forty-eight percent of all the ICU cohort had a positive IgG aCL test (Table 1); interestingly, fewer patients had elevated titers of IgM aCL (n=9, 21%), with only 2 patients having IgM without IgG. Although more COVID^+^ had aCL antibodies, the difference was not statistically significant. Longitudinally testing for anti-β2-GP1 and anti-PS/PT for IgG and IgM, as well as domain 1 β2-GP1 IgG revealed only one patient (COVID^+^) with positive serology for any of these autoantibodies. This patient seroconverted to IgM anti-PS/PT at days 5-7 of ICU hospitalization. Table 2 shows the temporal development of the aCL IgG and IgM antibodies stratified by COVID status. Late appearing (beyond 10 days after admission) aCL antibodies were not included in the statistical analyses to avoid survival and availability bias. Anti-CL were not associated with age or sex (not shown).

**Table 2:**
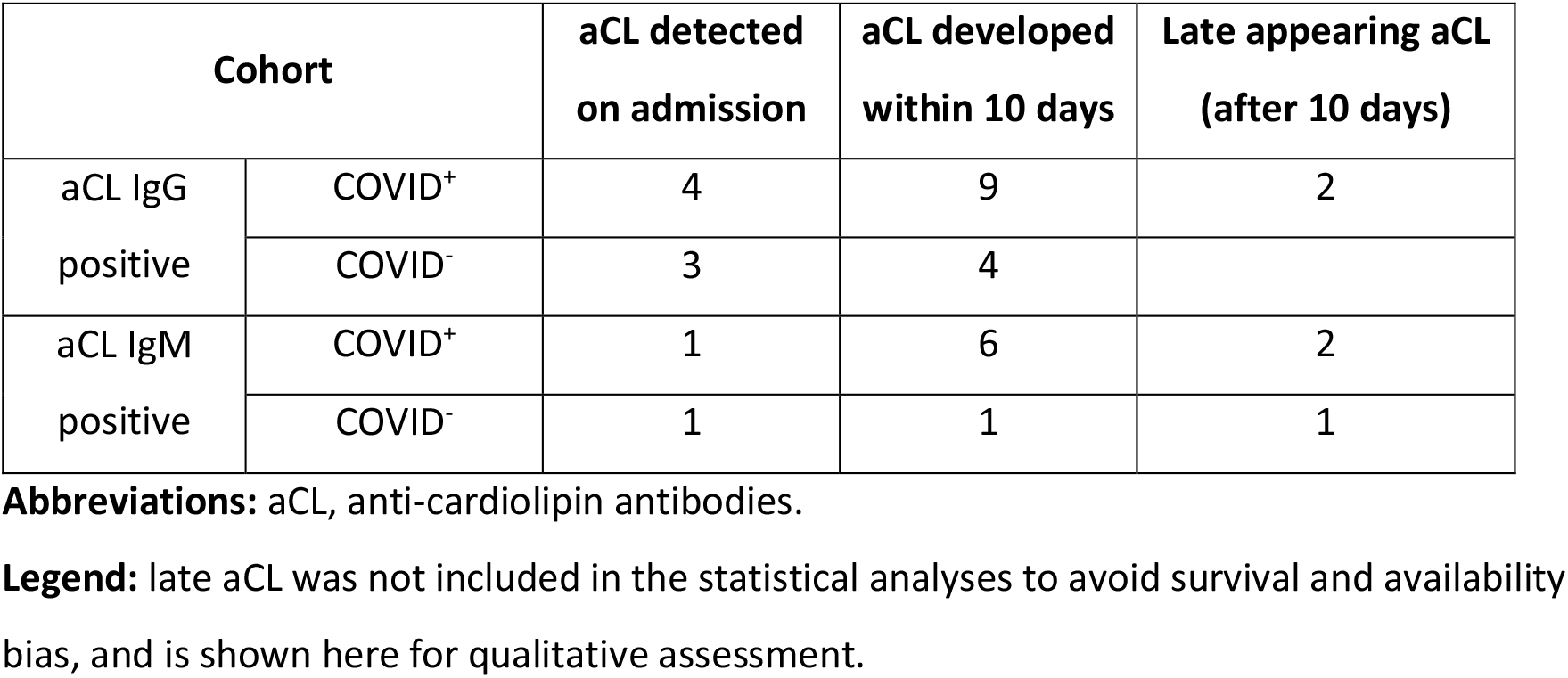
Development of aCL IgG and IgM over time.

| Cohort |  | aCL detected on admission | aCL developed within 10 days | Late appearing aCL (after 10 days) |
| --- | --- | --- | --- | --- |
| aCL IgG positive | COVID<sup>+</sup> | 4 | 9 | 2 |
|  | COVID<sup>-</sup> | 3 | 4 |  |
| aCL IgM positive | COVID<sup>+</sup> | 1 | 6 | 2 |
|  | COVID<sup>-</sup> | 1 | 1 | 1 |

**Abbreviations:** aCL, anti-cardiolipin antibodies.
**Legend:** late aCL was not included in the statistical analyses to avoid survival and availability bias, and is shown here for qualitative assessment.

### aCL vs disease severity, platelet counts and need for anticoagulation

Patients positive for aCL IgG demonstrated a consistent trend for worse outcomes in all the measures tested but this did not reach statistical significance after adjusting for multiple comparisons (Table 3). These trends remained when analyzed separately for COVID^+^ and COVID^-^ (not shown). Anti-CL IgG positive patients showed no significant differences in platelet counts, platelet to neutrophil ratio or the need for therapeutic anticoagulation (Table 3).

**Table 3:** **Association between aCL IgG and disease severity, platelet counts and need for anticoagulation**

|  | Cohort | All | aCL IgG positive | aCL IgG negative |
| --- | --- | --- | --- | --- |
|  | N | 42 | 20 | 22 |
| Age | Mean (CI) | 58.2 (62.7-54.1) | 55.9 (62.9-49) | 60.7 (66.4-55) |
| Sex | % male | 29/42 (69%) | 13/20 (65%) | 16/22 (73%) |
| Days from symptom onset to ICU | Mean (CI) | 6 (8.3-3.7) | 8.7 (12.8-4.6) | 3.4 (5.4-1.5) |
| APACHE II on ICU admission | Mean (CI) | 25.3 (27.6-22.9) | 25.7 (28.5-22.9) | 24.9 (28.8-20.9) |
| Mean of SOFA score for 1st 3 days | Mean (CI) | 9.6 (10.7-8.5) | 10.6 (12.2-9.1) | 8.7 (10.3-7) |
| Mean of SOFA score for 1st 7 days | Mean (CI) | 8.9 (10.1-7.8) | 10 (11.7-8.4) | 8 (9.5-6.4) |
| ICU days (censored) | Mean (CI) | 14.1 (17.3-10.8) | 16.6 (21.9-11.3) | 12.1 (16.5-7.6) |
| Death in ICU | N (%) | 13/42 (31%) | 8/20 (40%) | 5/22 (23%) |
| Mechanical ventilation days (censored) | Mean (CI) | 14.4 (18.9-10) | 18.2 (25.5-10.8) | 11.1 (16.4-5.7) |
| Total days of ventilation rescue measures | Mean (CI) | 2.9 (4.3-1.4) | 3.6 (5.6-1.5) | 2.3 (4.4-0.1) |
| Therapeutic anticoagulation used | N (%) | 8 | 4/20 (20%) | 4/22 (18%) |
| Mean platelet count | Mean (CI) | 239 (269-209) | 268 (321-216) | 212 (246-179) |
| Mean platelet to neutrophil ratio | Mean (CI) | 35.2 (42-28.4) | 34.8 (45.2-24.3) | 35.6 (45.4-28.9) |

**Abbreviations:** aCL, anticardiolipin; APACHE, Acute Physiology and Chronic Health Evaluation (score); ICU, intensive care unit; SOFA, sequential organ failure assessment (score).

### aCL association with ANA, antigen-specific autoantibodies and anti-cytokine autoantibodies

Although aCL IgG positivity was not associated with the presence of HEp-2 IFA ANA at a dilution of 1:160, it was significantly associated with higher ANA titers (p= 0.02), and this trend remained when analyzing the COVID^+^ and COVID^-^ patients separately. IgG aCL positivity was also significantly associated with anti-cytokine autoantibodies, both when analyzed for positive or high-positive anti-cytokine titers (p=0.003 for both, adjusted for multiple comparisons); this was not related to any particular anti-cytokine autoantibody, although anti-interferon-γ, anti-IL10 and anti-IL-17f were the most prevalent (supplemental table 2). When analyzing the aCL IgG positive according to their COVID-19 status, the COVID^+^ had significantly higher levels of anti-cytokine autoantibodies than the COVID^-^ (supplemental Table 3). Anti-CL IgG was not associated with antigen-specific autoantibodies, including SLE and myositis-related autoantibodies (not shown).

## DISCUSSION

The key observation of this study is that patients with positive IgG aCL showed a trend toward more severe disease regardless of whether they were COVID^+^ and COVID^-^. That is, while COVID^+^ patients showed non-significant trends towards worse respiratory outcomes when compared to COVID^-^, aCL status had an independent association with disease severity, and did not modulate the outcomes differentially based on COVID status. The pathological significance of aCL seropositivity is unclear since there were no major differences in platelet counts or thrombotic events in the two cohorts. Others have reported a high prevalence of aCL autoantibodies among COVID^+^ patients, but these studies lacked contemporaneous COVID^-^ control groups of similar disease severity^5 6 16 17^.

Although aCL tended to associate with COVID^+^, they did not associate with the presence of other antigen-specific autoantibodies, although they had a strong association with certain anti-cytokine autoantibodies. Interestingly, some patients had positive IgG aCL serology on ICU admission (Table 2) in the absence of another relevant comorbidity such as APS or SLE (supplementary table 1). These observations suggest that aCL positivity in the setting of acute severe respiratory illness may be a marker of a unique phenotype with variable temporal expression of aCL and anti-cytokine antibodies. The temporal dynamic is evidenced by the relatively long timeframe from symptom onset to ICU admission to the development of IgG aCL (Table 3). Our findings highlight the importance of longitudinal monitoring of acutely ill patients. It seems plausible that disparate conclusions in the literature with respect to the significance of APLAs in COVID-19 may relate to arbitrary sampling times and lack of longitudinal follow up in the setting of dynamic inflammatory diseases.

While some reports have included lupus anticoagulant (LAC) in their analyses, we did not because LAC is known to be an unreliable biomarker in severe illnesses where, C-reactive protein, anti-coagulant use and other factors confound its detection^18 19^. In this study, we used the anti-PS/PT test regarded by some as a surrogate for LAC (reviewed in^3^). However, only one patient developed anti-PS/PT 5-7 days after admission. Further, our observation that no patient had antibodies to β2-GP1 (an APS criteria antibody) or to domain 1 β2-GPI (reportedly higher specificity for APS) argues against the presence of APS in our cohort. In a study of 37 COVID+ acute respiratory disease versus 31 pre-pandemic (not contemporaneous) acute respiratory disease controls using a sample collected within 48 hours of admission, Frapard, et al. reported that 37 COVID patients exhibited more thrombotic events as compared to 31 pre-pandemic controls but the occurrence of APLA in the two groups was similar ^20^. Using APLA assays similar to ours, Borghi, *et al*. reported a low prevalence of APLA in COVID^+^ sera, where the most common target was IgG β2-GP1 (15.6%)^17^. In addition, the primary β2GP1 antibody targets were in domains 2-4 which are less specific for APS^17^. In agreement with our study, Bertin *et al*.^12^ and Borghi *et al*.^17^ concluded that APLA were not associated with major thrombotic events.

The main limitation of our study is the small sample size. The strengths of our study include its prospective, contemporaneous COVID^-^ cohort with similar severity of disease. Importantly, we tested a broad APLA serological panel longitudinally, providing a more robust assessment of its true prevalence and incidence than in other reported studies; this is particularly relevant for such acutely ill patients with dynamic clinical courses. Finally, our use of an extensive serological panel allowed us to better characterize the broad phenotype associated with aCL.

## Data Availability

The datasets used and/or analyzed during the current study are available from the corresponding author on reasonable request.

## DECLARATIONS

### Ethics approval and consent to participate

- This research was approved by Research Ethics Boards at St Michael’s Hospital and performed in accordance with the Helsinki Declaration of 1975 as revised in 2013.

### Consent for publication

- Not applicable

### Availability of data and materials

- The datasets used and/or analysed during the current study are available from the corresponding author on reasonable request.

### Competing interests

- MJF is the Director of MitogenDx. MJF is a consultant for and received speaking honoraria from Inova Diagnostics Inc (San Diego, CA) and Werfen International (Barcelona, Spain). All the other authors have no disclosures to declare.

### Funding

- St Michael’s Hospital Foundation, internal competitive grant to AB and CDS.
- Autoantibody testing was provided as a gift in kind by MitogenDx (Calgary, AB, Canada)

### Authors’ contributions

- This report is part of the COLOBILI study (Coronavirus longitudinal biomarkers in lung injury). AB and CDS are principal investigators; MJF, RR and AS are collaborators/co-investigators and UT is the research lead.
- RR, MJF, UT, and CDS conceived of the study; MJF, UT and RR wrote the manuscript drafts; AS, AB and CDS provided critique and technical guidance; UT performed the data analysis and creation of the figures. All authors edited the manuscript, through to the final version, read and approved the final submission.

## Acknowledgements

- The authors acknowledge the technical assistance of Haiyan Hou, Meifeng Zhang and Emily Walker in the MitogenDx Laboratory at the University of Calgary. We thank Marlene Santos, Gyan Sadhu, Imrana Khalid, and Sebastian Duncan, the research coordinators at St Michael’s Hospital Critical Care Research Unit. We are grateful to patients and families that have generously consented to the study.

## Study registration

- NCT04747782 – clinicaltrials.gov

## Patient and Public Involvement

- Patients and public were not involved in the design of the study. During the initial phases of the study, we obtained feedback from the patients and their substitute decision makers. Their concerns, questions and preferences were incorporated into improved processes for consent and collection of biological samples. The consent forms have checkboxes with optional aspects of the study, to accommodate different patient preferences. The results of the study will be disseminated in lay versions by St. Michael’s Hospital public relations and communications departments for the benefit of the public.

## Notes

### Clinical Trial

NCT04747782

### Funding Statement

St Michaels Hospital Foundation, internal competitive grant to AB and CDS.
Autoantibody testing was provided as a gift in kind by MitogenDx (Calgary, AB, Canada)

### Author Declarations

This research was approved by Research Ethics Boards at St Michaels Hospital and performed in accordance with the Helsinki Declaration of 1975 as revised in 2013.

