## Supplemental Methods for "Anti-cardiolipin and other anti-phospholipid antibodies in critically ill COVID-19 positive and negative patients": Suppl methods.pdf

### Online materials and methods

**Study design.** This report is part of the COLOBILI study – Coronavirus Longitudinal Biomarkers in Lung Injury, being conducted at St. Michael's Hospital (Toronto, ON, Canada). This is an observational cohort study that includes analysis of biological samples. The study was approved by the Research Ethics Board of St. Michael's Hospital (REB# 20-078). The inclusion criteria were all patients above age 18 years admitted to the Medical-Surgical or Trauma-Neuro intensive care units (ICU) with acute respiratory distress, suspected to have COVID-19. COVID-19 status was determined according to diagnostic PCR of nasopharyngeal swabs and/or endotracheal aspirates as described in detail below. The exclusion criteria were refusal to participate, inability to ascertain mortality status during the first 2 weeks of the study, failure to obtain a blood sample on either day 0 or 1, or individuals known to have had COVID-19 in the 4 weeks prior to admission in any setting. Patients were followed for up to 3 months in hospital or hospital discharge, whichever occurred first. The primary outcome was death in the ICU; secondary outcomes included death outside the ICU, ICU utilization metrics, and organ dysfunction measures and scores. Clinical data and blood samples were collected longitudinally immediately upon admission, as available, defined as day 0, and on the morning of days 1, 3, 5, 7 and 10; after day 10 or ICU discharge, they were sampled every 2 weeks. The study started on March 26th, 2020, and the first patient was recruited on March 29th, 2020. The study is ongoing; the last patient from the cohort presented in this manuscript was recruited on May 17th, 2020, and the data was censored for analysis on May 31st, 2020. No COVID-19 treatments were given to the patients beyond the standard of care since at the time there was no evidence of efficacy for any such treatments. Informed consent was obtained from the patients or their legal representatives; in case that was not possible, the patients were enrolled using a deferred consent model and kept in the study until they regained capacity, or a surrogate decision maker was identified.

**Data and sample collection.** Demographics, clinical data and clinical laboratory were collected from the patients' paper and electronic medical records, with auditing performed reciprocally by research coordination team members and curated by UT. To standardize handling and processing, blood samples were collected in EDTA tubes between 8:00 and 12:00 AM and kept on ice for up to 60 minutes until their processing in a dedicated translational research station located inside the ICU. They were then immediately frozen at -20 $^0$C on site, and transferred to -80 $^0$C for storage within 48 hrs. All procedures were performed by dedicated research personnel. Nasopharyngeal samples were obtained from all patients by bedside nurses and analyzed by the clinical laboratory using either the Altona RealStar SARS-CoV-2 RT-PCR Kit 1.0 or Cepheid GeneXpert Xpert Xpress SARS-CoV-2 assay. Endotracheal tube aspirates were analyzed using the Seegene Allplex 2019-CoV Assay. All patients had a nasopharyngeal PCR performed; intubated patients had an endotracheal aspirate sent as well. Further PCR tests were repeated by the clinical and infection control teams at their discretion if there was suspicion of a false negative result based on clinical observations or to confirm negativity. All patients in the PCR negative cohort had at least two negative tests performed acutely, except one patient who had only one test done acutely. To analyze longitudinal trends, only patients with 3 or more longitudinal sampling times were included in the study. To mitigate bias, five patients with shorter ICU admissions were included; 2 had early deaths and 3 had early discharges.

**Experimental procedures.** Plasma samples were stored and managed under a standard operating procedure which included shipping on dry ice and storage at -80C until assay performance by Mitogen Diagnostics Laboratory (MitogenDx, Calgary, AB, Canada). Anti-cardiolipin, anti-β2-GP1 and anti-PS/PT complex were tested for IgG and IgM, as well as IgG anti-domain 1 β2-GP1 – all by ELISA and chemiluminescence (Inova Diagnostics, San Diego, CA USA). A HEp-2 indirect immunofluorescence assay (IFA) was used to detect anti-cellular antibodies (also referred to as anti-nuclear antibodies (ANA) – see "nomenclature" below)[1] (NOVA Lite HEp-2, Inova Diagnostics, San Diego, CA) at a serum dilution of 1:80 and read on an automated instrument (Nova View, Inova Diagnostics) which interpolates fluorescence intensity to an end point titer[2]. IFA staining patterns were classified according to the International Consensus on Autoantibody Patterns (ICAP, https://anapatterns.org/index.php)[3], and considered positive at a dilution ≥1:160. All samples were also tested for systemic autoimmune disease-related autoantibodies by a FIDIS Connective13 addressable laser bead immunoassay (ALBIA) (TheraDiag, Paris, France) detecting antibodies to Sm/U2-U6 ribonucleoprotein (RNP), U1-RNP, SSA/Ro60, SSB/La, Ro52/Tripartite Motif Protein 21 (TRIM21), histones, and ribosomal P, read on a Luminex 200 system using the MLX-Booster software. A cut-off of >40 units was considered positive. Anti-dsDNA positivity and titers were detected by a chemiluminescence test (Inova Diagnostics, San Diego, USA). A cut-off of <27 chemiluminescence units was considered within normal range, 27-35 was indeterminate, and >35 was positive. All samples were also tested for autoantibodies associated with autoimmune inflammatory myopathies using a multiplexed solid phase line immunoassay: Ro-52/TRIM21, OJ, EJ, PL-12, PL-7, SRP, Jo-1, PM-75, PM-100, Ku, SAE1, NXP2, MDA5, TIF1γ, Mi-2α, Mi-2β (Euroimmun AG, Luebeck, Germany), and anti-NT5c1A by ALBIA[4]. The following anti-cytokine antibodies were assayed using an ALBIA (Millipore, Oakville, ON, Canada; HCYTAAB-17K-15) read on a Luminex 200 system: BAFF, GMCSF, IFN-β, IFN-γ, IL-1a, IL-6, IL-8, IL-10, IL-12p40, IL-15, IL-17a, IL-17f, IL-18, IL-22 and TNF-α. The manufacturer's thresholds were 500 for positive and 1000 for high-positive (arbitrary units). All tests were performed according to the manufacturer's instructions.

**Nomenclature.** There is considerable heterogeneity in the nomenclature of autoimmune assays in the literature and clinical practice; therefore, we used the most contemporary usages. Autoantibodies is a general term that encompasses all of the autoimmune humoral responses assayed. The HEp-2 IFA, although including anti-cytoplasmic and anti-mitotic cell antibodies, are commonly referred as anti-nuclear antibodies (ANA), and we have adopted that usage for clarity. The AAB test results that identified specific, named antigens (see details above), were called collectively antigen-specific autoantibodies. We have further separated them into myositis-related and non-myositis-related AAB. Anti-cytokine autoantibodies are referred to directly.

**Data analysis.** All the data was organized and analyzed by UT. The data was censored on May 31st, 2020; only 5 patients had censored data for the primary outcome, death in the ICU within 3 months. Given the elapsed time until censoring, the risk of right-censoring bias is low. ANOVA was used for continuous variables and Fisher's exact test was used for categorical variables at $\alpha=0.05$, adjusted for multiple comparisons as indicated in the text using the false discovery rate at $q=0.05$. All statistical and graphical analyses were performed on JMP Pro (version 15.2.1; SAS Institute Inc, Cary, NC, USA).
