## Supplemental Tables for "Anti-cardiolipin and other anti-phospholipid antibodies in critically ill COVID-19 positive and negative patients": Supplemental tables.pdf

**Supplemental Table 1: Premorbid Clinical Characteristics and Therapeutics**

| | All (N) | COVID+ (N) | COVID- (N) |
|---|---|---|---|
| | 42 | 22 | 20 |
| Respiratory PMH | 18 | 8 | 10 |
| Cardiovascular PMH | 19 | 11 | 8 |
| Renal PMH | 7 | 6 | 1 |
| Type 2 Diabetes | 20 | 12 | 8 |
| Hypertension | 24 | 14 | 10 |
| Other comorbidities | 37 | 18 | 19 |
| Premorbid steroid used | 3 | 1 | 2 |
| Premorbid immunomodulatory medication use | 2 | 1 | 1 |
| Premorbid ACEi/ARB use | 15 | 10 | 5 |

Abbreviations: ACEi, Angiotensin-converting enzyme inhibitors; ARB, angiotensin receptor blocker; PMH, past medical history.

Legend: "Other comorbidities" include autoimmune diseases: Myasthenia gravis among COVID+, and autoimmune hemolytic anemia, rheumatoid arthritis and multiple sclerosis among the COVID-. No statistically significant difference between COVID+ and COVID- patients for all variables were detected using ANOVA for continuous variables and Fisher's exact test for categorical variables at $\alpha=0.05$

**Supplemental Table 2: Association between aCL IgG and high-titre anti-cytokine autoantibodies**

| | **Cohort** | **All** | **aCL positive** | **aCL negative** |
|---|---|---|---|---|
| | **N** | 42 | 20 | 22 |
| ALL | *N (%)* | 16/42 (38%) | 13/20 (65%) | 3/22 (14%) |
| anti-GMCSF | *N (%)* | 1/42 (2%) | 1/20 (5%) | 0/22 (0%) |
| anti-IFN-γ | *N (%)* | 7/42 (17%) | 6/20 (30%) | 1/22 (5%) |
| anti-IL-1a | *N (%)* | 1/42 (2%) | 0/20 (0%) | 1/22 (5%) |
| anti-IL-6 | *N (%)* | 5/42 (12%) | 3/20 (15%) | 2/22 (9%) |
| anti-IL-10 | *N (%)* | 5/42 (12%) | 5/20 (25%) | 0/22 (0%) |
| anti-IL-12p40 | *N (%)* | 2/42 (5%) | 2/20 (10%) | 0/22 (0%) |
| anti-IL-17a | *N (%)* | 2/42 (5%) | 2/20 (10%) | 0/22 (0%) |
| anti-IL-17f | *N (%)* | 4/42 (10%) | 4/20 (20%) | 0/22 (0%) |
| anti-IL-22 | *N (%)* | 1/42 (2%) | 0/22 (0%) | 1/22 (5%) |

Abbreviations: aCL, anticardiolipin; GMCSF, granulocyte-macrophage colony-stimulating factor; IFN, interferon; IL, interleukin.

Legend: The table reports the number of patients with high titres of anti-cytokine antibodies at any time during the first 10 days of ICU admission. "ALL" represents all patient having a high titre of anti-cytokine antibody of any type during that period. The numbers of the specific anti-cytokine antibodies sum to more than "ALL" since some patients had more than one high titre anti-cytokine antibody. Once adjusted for multiple comparisons, there were no statistically significant differences between aCL positive and aC negative- for any of the results (Fisher's exact test at α=0.05 followed by the false discovery rate at q=0.05). The following anti-cytokine AAB did not show high levels in any of the patients: anti-BAFF, anti-IFN-β, anti-TNF-α, anti-IL8, anti-IL-15 and anti-IL-18.

**Supplemental Table 3: Association between aCL IgG and anti-cytokine autoantibodies per COVID-19 status**

| | | | Anti-cytokine autoantibody titers | |
|---|---|---|---|---|
| | | | **Positive** | **High-positive** |
| COVID$^+$ | aCL IgG positive | *N, %* | 12/13, 92%* | 9/13, 69% |
| COVID$^-$ | | | 4/7, 57% | 4/7, 57% |

Abbreviations: aCL, anti-cardiolipin antibodies.

Legend: The asterisk (*) represents a significant association between aCL IgG and anti-cytokine autoantibodies (Fisher's exact test, p=0.006, adjusted for multiple comparisons).
